# Patterns of Medication Use and Prescription Fills for Cardioprotective Anti-Hyperglycemic Agents in the United States

**DOI:** 10.1101/2022.01.31.22270187

**Authors:** Arash A Nargesi, Callahan Clark, Lian Chen, Mengni Liu, Abraham Reddy, Samuel Amodeo, Evangelos K Oikonomou, Marc A Suchard, Darren K McGuire, Zhenqiu Lin, Silvio Inzucchi, Rohan Khera

## Abstract

**Importance:** Selected glucagon-like peptide-1 receptor agonists (GLP-1RAs) and sodium glucose cotransporter-2 inhibitors (SGLT2i) have cardioprotective effects in patients with type 2 diabetes and elevated cardiovascular risk. Prescription of these agents by clinicians and their consistent use by patients are essential to realize their benefits.

**Objective:** To assess the patterns of use and prescription fills of GLP-1RAs and SGLT-2i.

**Design:** Cross-sectional for medication use and prospective for prescription fills in 2018-2020

**Setting:** Nationwide de-identified US administrative claims database of Medicare Advantage and commercially insured adults.

**Participants:** Individuals 18 years of age and older with type 2 diabetes

**Exposures:** Comorbidities representing guideline-directed indications of atherosclerotic cardiovascular disease (ASCVD) for GLP-1RAs, and ASCVD, heart failure, and diabetic nephropathy for SGLT2i.

**Main Outcomes and Measures:** Medication use and monthly fill rates for 12 months following initiation of therapy by calculating the proportion of days with consistent medication use.

**Results:** Among 587,657 individuals with type 2 diabetes, 80,196 (13.6%) were prescribed GLP-1RAs and 68,149 (11.5%) SGLT2i during 2018-2020. This represented 12.9% and 10.5% of individuals with indications for each medication, respectively. Based on monthly counts of new prescriptions, there were no changes in the uptake of either drug class during 2019-2020. Among new initiators, fill rate was 52.5% for GLP-1RAs and 52.9% for SGLT2i one year after initiation. One-year fill rates were higher for patients with commercial insurance than those with Medicare Advantage plans for both GLP-1RAs (59.3% vs 51.0%, p-value<0.001) and SGLT2i (63.4% vs 50.3%, p-value<0.001). After adjusting for comorbidity profile, there were higher prescription fills for patients with commercial insurance (versus Medicare Advantage, OR 1.17, 95% CI [1.06-1.29] for GLP-1RAs, and 1.59 [1.42-1.77] for SGLT2i); and higher income (top quartile versus others, OR 1.09 [1.06-1.12] for GLP-1RAs, and 1.06 [1.03-1.10] for SGLT2i).

**Conclusions and Relevance:** In 2018-2020, use of GLP-1RAs and SGLT2i remained limited to fewer than 1 in 8 individuals with type 2 diabetes meeting criteria for evidence-based guideline and professional society recommendations, with one-year fill rates around 50%. The low and inconsistent use of these medications compromises their longitudinal health outcomes benefits in a period of expanding indications for their use.

## BACKGROUND

Glucagon-like peptide-1 receptor agonists (GLP-1 RAs) and sodium glucose cotransporter-2 inhibitors (SGLT-2i) are recommended in clinical practice guidelines for treatment of patients with type 2 diabetes and compelling cardiovascular and kidney indications, independent of glucose control.^1-5^ This is underpinned by the results from seminal, large outcome trials in patients with type 2 diabetes reported between 2015 and 2019.^6,7^ Guidelines and professional society recommendations began to endorse their use as early as in 2017, with consistent Class I recommendations beginning in 2018, with omission of all glucose control considerations harmonious across guidelines and society recommendations by 2020. Both drug classes confer protection against major atherosclerosis-based adverse cardiovascular events in patients with atherosclerotic cardiovascular disease (ASVCD).^8^ SGLT-2i also lower the risk of hospitalization for heart failure and prevent worsening kidney function in patients with type 2 diabetes and ASCVD risk or with diabetic kidney disease.^6^

Despite the expanding indications for GLP-1 RAs and SGLT-2i in clinical guidelines and society recommendations for cardiovascular and kidney benefits, the overall prescriptions of these medications remain low.^9-12^ Furthermore, the clinical benefits of GLP-1 RAs and SGLT-2i are only achieved if patients continually take these medications after prescription, underscoring the importance of both prescription and consistent use. In the absence of high-quality data on the evolving use of GLP-1 RAs and SGLT-2i in the real world, assessment of their consistent use after initiation of therapy has been challenging. In addition, affordability of these agents remains a major concern, potentially affecting both their initiation and continuation of use.

In this US national study, we evaluated the contemporary patterns of prescription for GLP-1 RAs and SGLT-2i among individuals with type 2 diabetes meeting evidence-based indication for their use and assessed the actual use of these agents by measuring their monthly fill rates after starting treatment. The patterns of use of these medications were evaluated across key patient subgroups defined by guideline- and professional-society recommended clinical indications for their use and the insurance coverage of treated individuals.

## METHODS

### Data Sources

We used Optum Labs’ de-identified administrative claims data, which contains longitudinal enrollment records, medical claims, and pharmacy claims for Medicare Advantage and commercially insured beneficiaries and represents a diverse population across the US.^13^ These records include information on patient demographics, type of insurance plan (Medicare Advantage vs commercial), healthcare conditions, and their treatments captured as standard health insurance claims.

### Primary Study Population

The study population included individuals 18 years of age or older with 36 months of continuous enrollment in Medicare Advantage with Part D coverage or commercial insurance with pharmacy coverage between January 2018 through December 2020 who had two or more claims more than 90 days apart with a principal or secondary diagnosis of type 2 diabetes (**eTable 1, online supplement)**.

The diagnosis of type 2 diabetes was defined based on International Classification of Disease tenth revision (ICD-10) codes. Antihyperglycemic treatment was defined as the receipt of one or more agents included in Standards of Medical Care in Diabetes by the American Diabetes Association.^14^ These included GLP-1 RAs, SGLT-2i, metformin, sulfonylureas, thiazolidinediones, dipeptidyl peptidase-4 (DPP-4) inhibitors, and insulins. The individual agents included in each drug class are presented in **eTable 2 (online supplement)**. Fixed-dose drug combinations were considered equivalent to taking the individual component medications separately. Drug information was obtained from pharmacy claims corresponding to a cumulative supply >30 days between January 1, 2018, and December 31, 2020. The Medicare Advantage and commercially insured enrollees in the study represented all individuals with available claims in the database who met the inclusion/exclusion criteria.

### Study Exposures

We used clinical practice recommendations of the American Diabetes Association (ADA), American College of Cardiology (ACC), and American Association of Clinical Endocrinologists and American College of Endocrinology (AACE/ACE) for management of type 2 diabetes to define indications for GLP-1 RAs and SGLT-2i. These indications included heart failure and diabetic nephropathy for SGLT-2i, and ASCVD for both SGLT-2i and GLP-1 RAs.^2,14,15^ The criteria for identifying these indications were consistent with prior studies^6,7^ and are detailed in the **eTable 3** (**online supplement**). **Figure 1** represents the timeline of data collection in the context of clinical trials and approval of cardiovascular and kidney indications in product labels for individual agents within each drug class.

**Figure 1.**
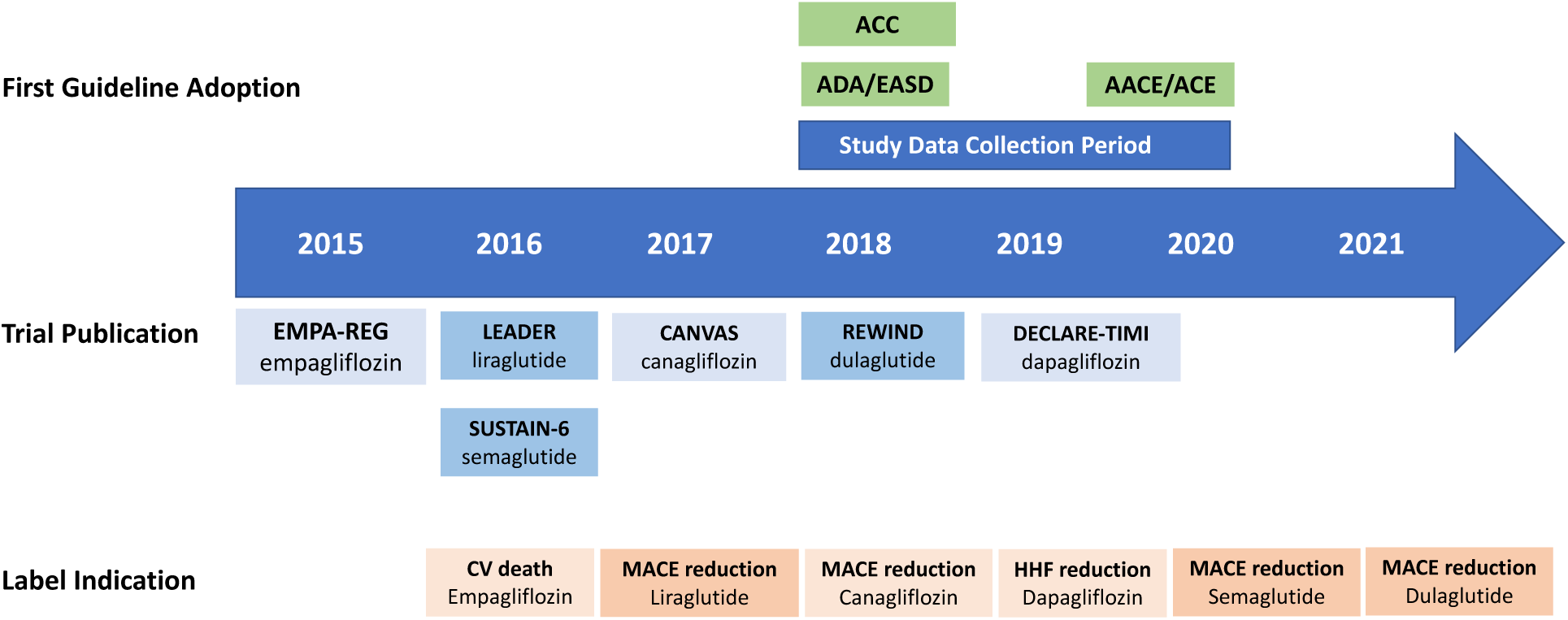
Timeline of data collection, approval of drug labels, and uptake by clinical practice guidelines for GLP-1 RAs and SGLT-2i. Figure represents the timeline of FDA approval for GLP-1RA and SGLT-2i drug labels, adoption by clinical practice guidelines, and data collection in this study. Abbreviations: ACC, American College of Cardiology; ADA, American Diabetes Association; EASD, European Association of the Study of Diabetes; AACE, American Association of Clinical Endocrinologists; ACE, American College of Endocrinology; CV, cardiovascular; MACE, major adverse cardiovascular event; HHF, hospitalization for heart failure.

Individuals with claim-based evidence of contraindications for each medication based on the US Food and Drug Administration (FDA) product information were considered non-eligible. These conditions included medullary thyroid carcinoma and multiple endocrine neoplasia syndrome type 2 for GLP-1 RAs, and chronic kidney disease (CKD) stage IV-V, end stage kidney disease (ESKD), and dialysis for SGLT-2i (**eTable 3, online supplement)**.

### Study Covariates

We included demographic characteristics of the study population, including age, sex, type of health insurance plan, neighborhood income (as identified by zip code), and comorbid conditions included in the Diabetes Complications and Severity Index (DCSI) and Charlson Comorbidity Index (CCI).^16,17^ In addition, the use of other cardiovascular therapies was identified within pharmacy claims, which included statins, beta blockers, angiotensin converting enzyme inhibitors (ACEI), angiotensin II receptor blocker (ARB), and oral anticoagulants (including warfarin and direct-acting oral anticoagulants). The characteristics of the study population and medication use were obtained from insurance claims in 2018 (the baseline study year).

### Study Outcomes

The study focused on two key outcomes: 1) proportionate prescription for GLP-1 RAs and SGLT-2i among individuals with type 2 diabetes, overall and among those with compelling indications; and 2) monthly fill rates for GLP-1 RAs and SGLT-2i for 12 months after initial prescription of the therapy.

The proportionate use of GLP-1 RAs and SGLT-2i were defined as the proportion of individuals with any prescription for these agents during the 3-year window from January 2018 through December 2020. We also assessed trends in new prescriptions for each drug class among eligible individuals between January 2019 through December 2020. New initiators were defined as individuals who were initially prescribed a GLP-1 RA and/or an SGLT2i after at least a 12-month period without any prescriptions for the respective medications. Data from January through December 2018 were used to ensure no prescription prior to starting the treatment in new initiators.

The monthly fill rates for GLP-1 RAs and SGLT-2i were assessed by calculating the proportion of days covered (PDC) among new initiators. PDC is defined as the proportion of days in a certain period of time with evidence of medication dispense based on pharmacy claims and is a measure of consistent fill of the medication.^18^ To ensure that each person had at least 12 months of claims to provide fill information, these analyses were restricted to new initiators in 2019, with the first fill occurring between January through December 2019, and the subsequent fill data derived from pharmacy claims through December 2020. Monthly fill rates for metformin and sulfonylureas were similarly assessed in new initiators of each drug class in the similar period to compare with GLP-1 RAs and SGLT-2i.

### Statistical Analysis

Categorical variables are presented as frequency and percentages, and continuous variables as mean and standard deviations or median and interquartile ranges, as appropriate. Differences in characteristics across categories of individuals based on indications for GLP-1 RAs and SGLT-2i were compared using chi square for categorical variables and analysis of variance for continuous variables.

Independent predictors of consistent fills for each drug class were tested using logistic regression. PDC for SGLT-2i and GLP-1 RAs were averaged among new initiators of each medication over 12 months after initiation of treatment and then dichotomized into a binary outcome variable, comparing the highest tertile with the lower two tertiles of PDC. Neighborhood income was also dichotomized into a binary variable, comparing top quartile with the lower three quartiles before inclusion in the regression model. Age, sex, neighborhood income, type of insurance plan, DCSI score, CCS score, ASCVD, heart failure, and diabetic nephropathy were considered as independent factors in the model.

Analyses were performed using R 3.4.0 (CRAN). All hypothesis tests were 2-sided, with a level of significance set at 0.05. The UnitedHealth Group Office of Human Research and the Yale Institutional Review Board exempted this study from review and waived informed consent as the study was limited to retrospective analyses of de-identified data and compliant with the Health Insurance Portability and Accountability Act (HIPAA) of 1996.

## RESULTS

### Characteristics of the Study Population

We identified 587,657 individuals with type 2 diabetes and 36 months of continuous enrollment. Demographic and clinical characteristics of study population across eligibility groups are represented in **Table 1**. The mean age of the study population was 72.8 (9.2) years, 52% were women, median neighborhood income was $53,317 [IQR $46,338-$62,553], 89% had Medicare Advantage coverage and 11% were commercially insured. The study population included 209,983 individuals with ASCVD (35.7%), 68,506 with heart failure (11.6%), and 159,436 with diabetic nephropathy (27.1%).

**Table 1.**
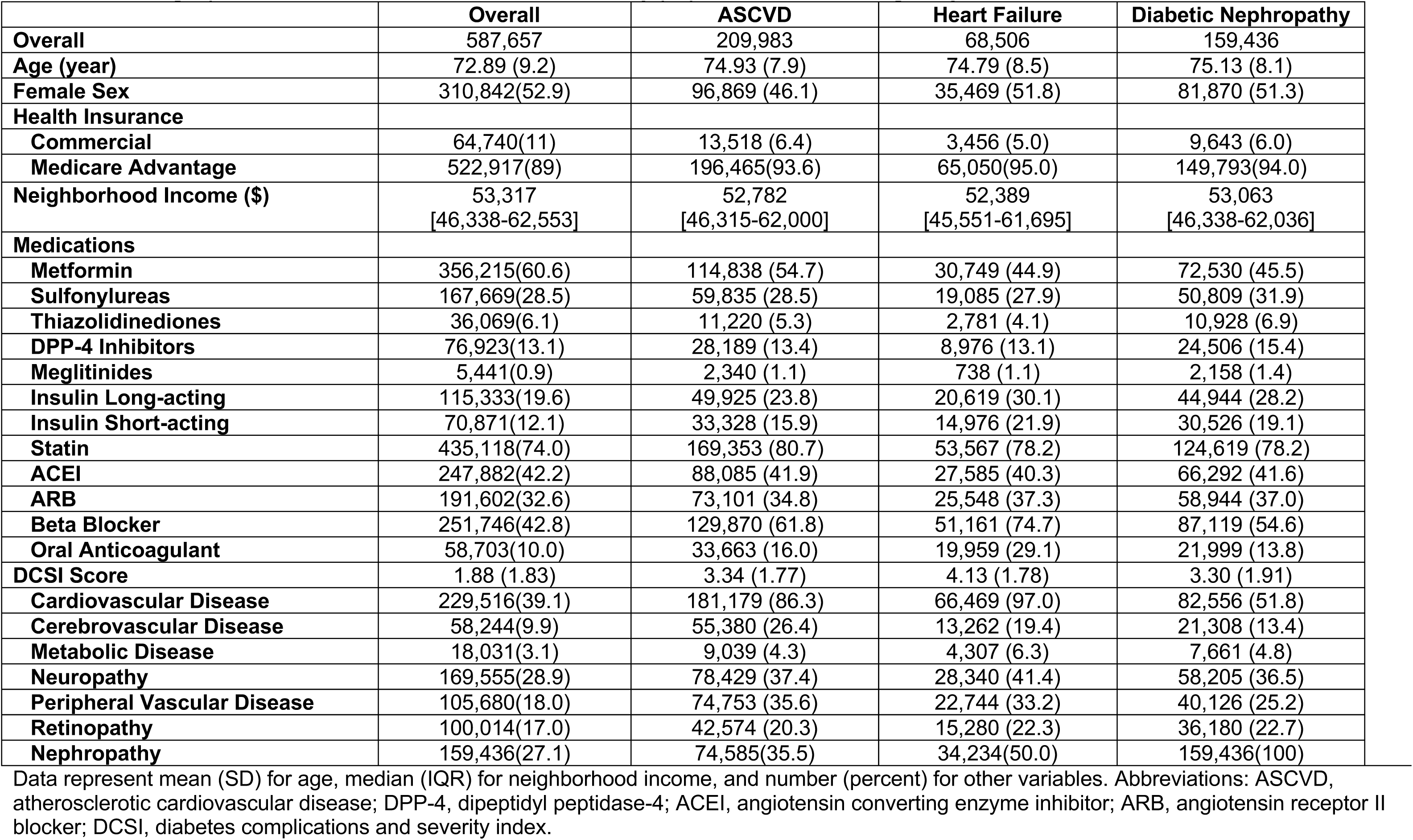
Demographic and clinical characteristics of study population across eligibility for GLP-1 RAs and SGLT-2i

### Use of GLP-1 RAs and SGLT2i

Overall, 80,196 individuals filled GLP-1 RA prescriptions in 2018-2020, representing 13.6% of individuals with type 2 diabetes and 12.9% of those with type 2 diabetes and coexisting established ASCVD. For SGLT-2i, a total of 68,149 individuals filled a prescription in this drug class, representing 11.5% of individuals with type 2 diabetes and 10.5% of those with type 2 diabetes and any indication, including 23,014 individuals with ASCVD (11%), 6,367 with heart failure (9.3%), and 14,224 with diabetic nephropathy (8.9%). The proportional use of GLP-1 RAs and SGLT-2i among individuals with indications are represented in **Figure 2**. During 2019-2020, there was no significant trend in new prescriptions for either GLP-1 RAs or SGLT-2i among eligible patients (**eFigure 1, online supplement)**. Demographic and clinical characteristics of individuals filling GLP-1 RAs and SGLT-2i are presented in **eTable 4 (online supplement)**.

**Figure 2.**
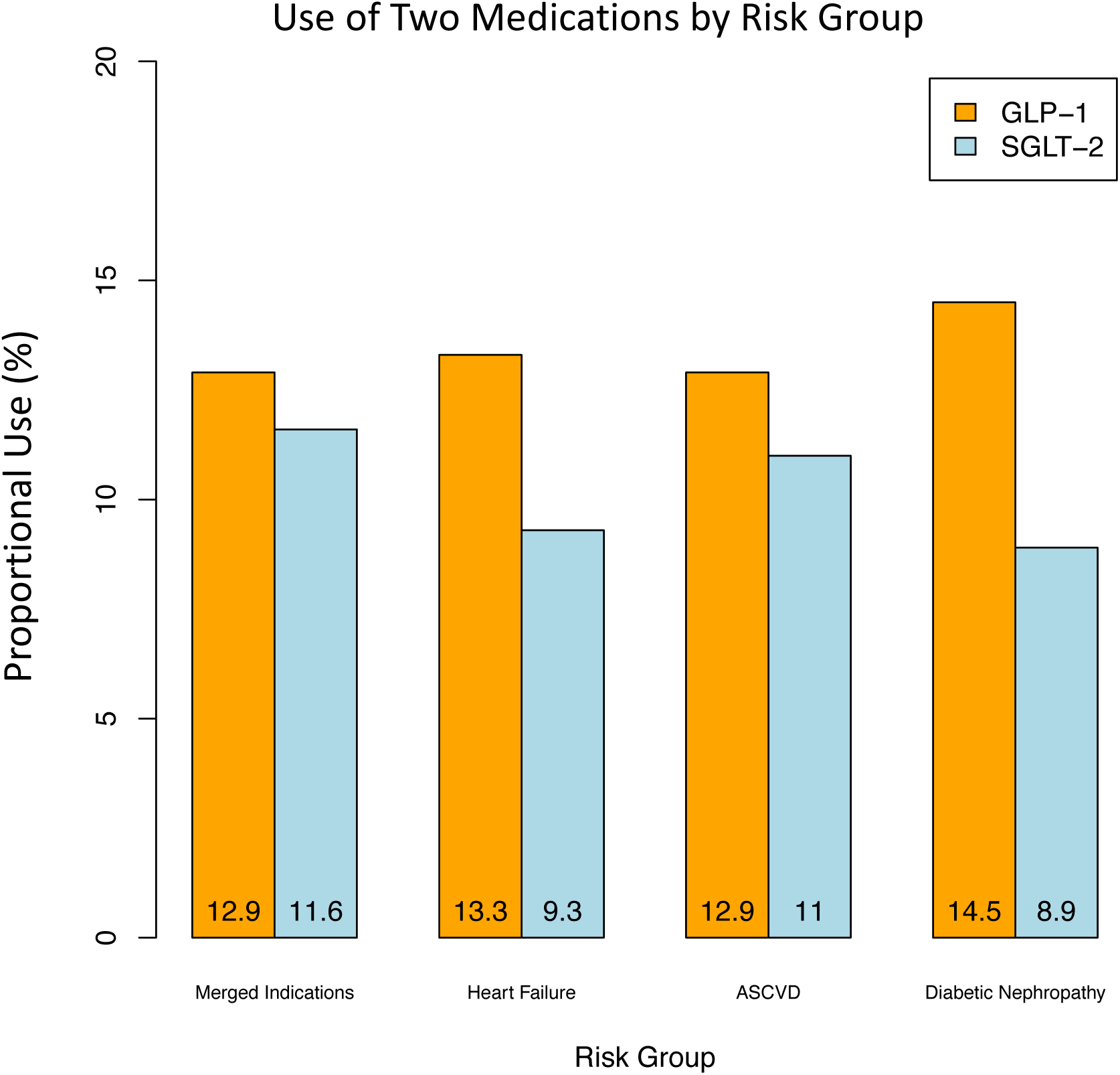
Use of GLP-1 RAs and SGLT-2i among eligible patients. Data represent percentage of individuals filling a prescription for each medication overall and in selected subgroups by indication. Indications for each drug class were evaluated independently based on guideline and professional society recommendations and included ASCVD for GLP-1 RAs, and ASCVD, heart failure, and diabetic nephropathy for SGLT-2i. Abbreviations: GLP-1; glucagon-like peptide-1 receptor agonist; SGLT-2, sodium glucose cotransporter 2 inhibitor; ASCVD, atherosclerotic cardiovascular disease.

### Monthly fill rates for GLP-1 RAs and SGLT-2i after initiation of therapy

Monthly fill rates for GLP-1 RA and SGLT-2i prescriptions after starting treatment are represented in **Figure 3**. Fill rate was 63.7% for GLP-1 RAs and 67.8% for SGLT-2i 3 months after initiation of therapy. For GLP-1 RAs, one-year fill rate was 52.5%, and for SGLT2i 52.9% after starting treatment. For GLP-1 RAs, one-year fill rate was 50.8% for patients with ASCVD. Across patient subgroups, one-year fill rates for SGLT2i were 51.5% for individuals with ASCVD, 52.7% for individuals with heart failure, and 49.8% for those with diabetic nephropathy (**eFigure 2, online supplement)**.

**Figure 3.**
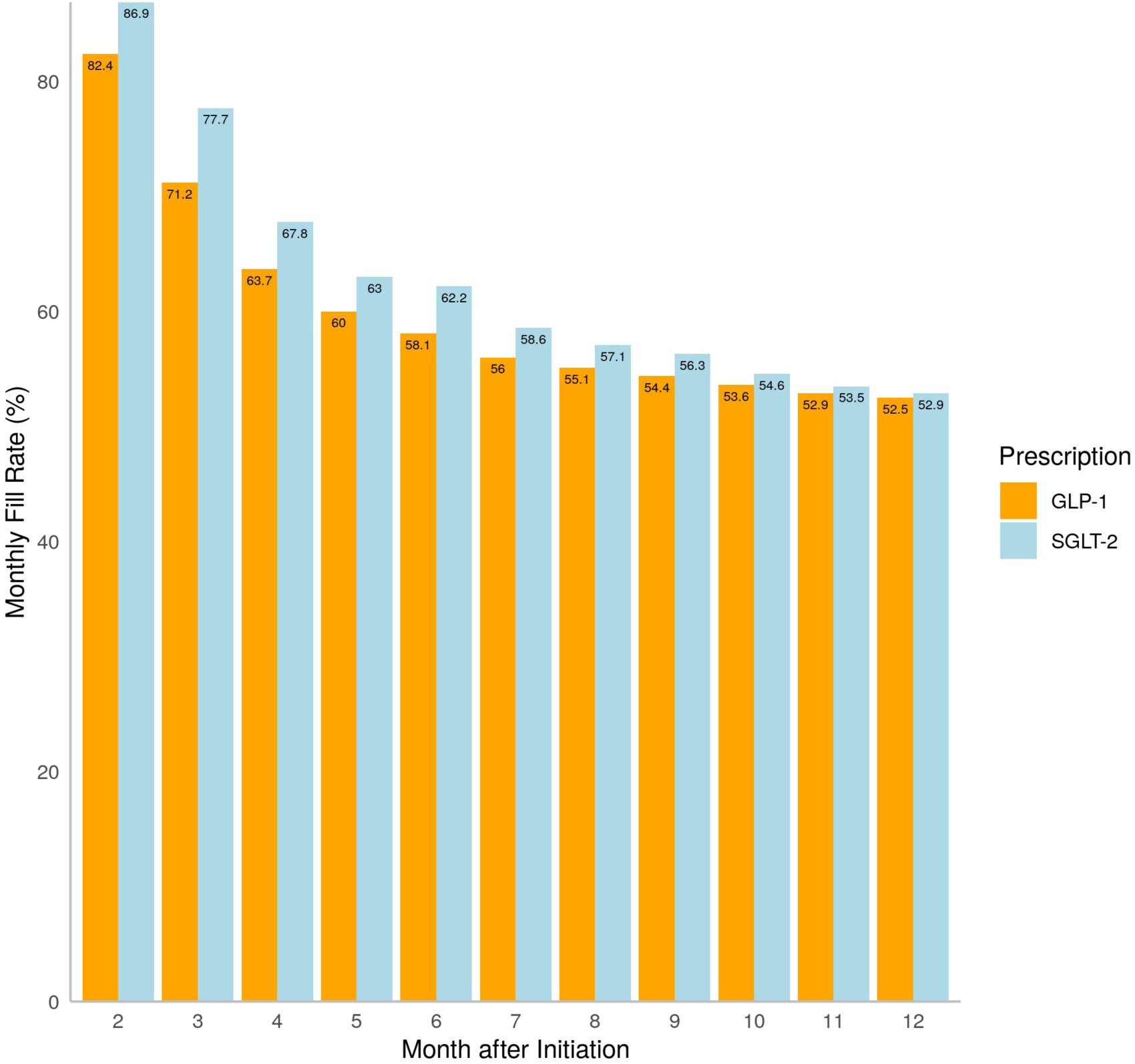
Monthly fill rates for GLP-1 RA and SGLT-2i prescriptions among new initiators in 2019-2020. Data represent monthly fill rates based on the proportion of days covered among new initiators of each drug class for 12 months after starting treatment. Abbreviations: GLP-1; glucagon-like peptide-1 receptor agonist; SGLT-2, sodium glucose cotransporter-2 inhibitor.

One-year fill rates were higher for individuals with commercial insurance than those with Medicare Advantage plans for GLP-1 RAs (59.3% vs 51.0%, p-value<0.001) and SGLT-2i (63.4% vs 50.3%, p-value<0.001). **eFigure 3** represents monthly fills for GLP-1 RA and SGLT-2i prescriptions in commercially insured and Medicare Advantage beneficiaries (**online supplement)**. One-year fill rates among new initiators were 55.8% for metformin and 62% for sulfonylureas, as represented in **eFigure 4 (online supplement)**.

### Predictors of consistent use of GLP-1 RAs and SGLT2i

In multivariable model with one-year fill as outcome, higher neighborhood income (comparing top income quartile with the other quartiles combined: OR 1.09 [95% CI: 1.06-1.12] for GLP-1 RAs; and 1.06 [1.03-1.1] for SGLT-2i) and commercial insurance (compared with Medicare Advantage: OR 1.17 [1.06-1.29] for GLP-1 RAs and 1.59 [1.42-1.77] for SGLT-2i) were associated with higher prescription fills. History of heart failure and diabetic nephropathy were not associated with fill rates for either GLP-1 RAs or SGLT-2i. Established ASCVD was associated with lower prescription fills for SGLT-2i (OR 0.88 [0.80-0.96]) (**Figure 4**).

**Figure 4.**
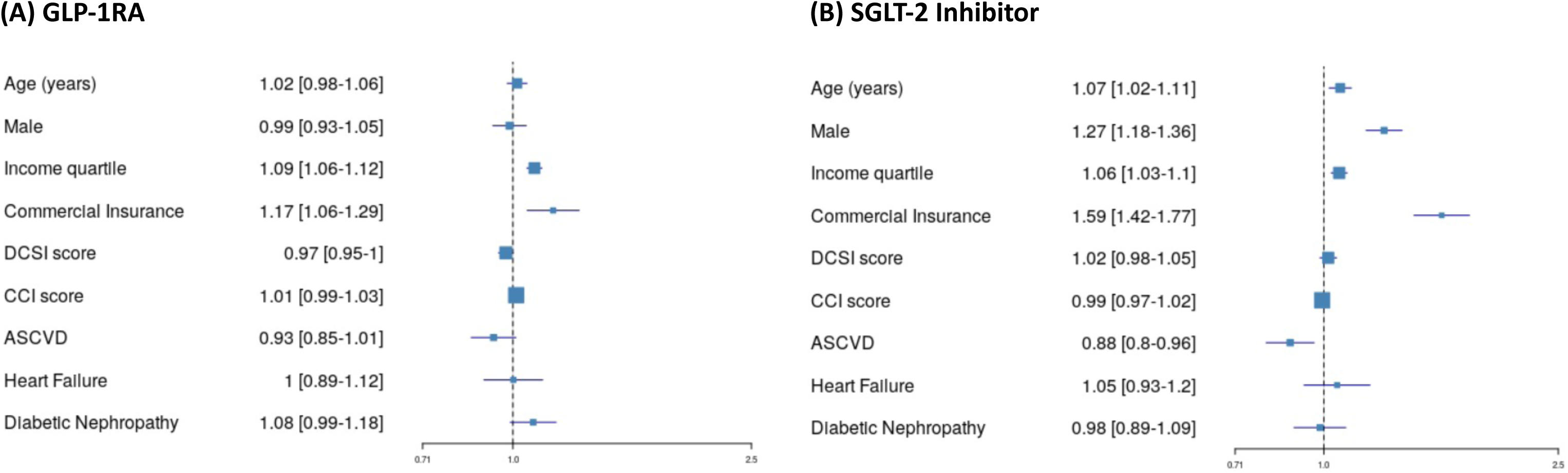
Multivariable predictors of one-year fill rates for GLP-1 RA and SGLT2i prescriptions. Data represent odds ratio (95% CI) in multivariable model with one-year prescription fill rates for GLP-1RA and SGLT-2i prescriptions as outcome. Abbreviations: GLP-1 RA; glucagon-like peptide-1 receptor agonist; SGLT-2, sodium glucose cotransporter-2; DCSI, diabetes complications and severity index; CCI, Charlson comorbidity index; ASCVD, atherosclerotic cardiovascular disease.

## DISCUSSION

In this nationwide study analyzing administrative data from the United States, GLP-1 RAs and SGLT-2i were used in 11 to 13 percent of adults with type 2 diabetes in 2018-2020, with only 1 in 8 individuals with clear clinical indications receiving prescriptions for these medications. The uptake of GLP-1 RAs and SGLT-2i remained unchanged in 2019-2020 with no significant trend in the counts of new prescriptions for these agents in this period. Furthermore, among patients who started these medications, only two-thirds were consistently taking these medications at 3 months and only half one year after starting the treatment. Patients with guideline-directed indications for the use of these medications had similarly low fill rates, with high income and coverage under commercial insurance associated with higher fills one year after initiation of therapy.

The uptake of the novel antihyperglycemic medications among patients with type 2 diabetes remains low through 2020 despite multiple clinical trials reporting cardiovascular and kidney protective effects of these agents since 2016, with endorsement in clinical practice guidelines and society recommendations since 2018. The fill rates of these medications are similarly low among patients with guideline-directed indications, with no significant increase in new prescriptions during 2019-2020. The present results support the observations of prior studies suggesting low rates of prescriptions of these medications,^9-11^ with this study reporting the latest data on the real-world use of GLP-1 RAs and SGLT-2i in a large and diverse population across the US. The constellation of these findings suggests a guideline-discordant prescription pattern that does not adequately incorporate trial-proven non-glycemic benefits of these two drug classes.

We found a month-to-month decline in the number of patients who actively fill these agents in the year following their initial prescription, such that at 12 months after initiation only half of the prescriptions were filled. Moreover, this pattern of underfilling of prescriptions for these agents did not differ among patients with and without risk for cardiovascular and kidney disease, suggesting a lack of selectivity or emphasis on their importance in clinical use. We also found evidence of a larger drop off in prescription fills for these agents compared with the older and less expensive medications, such as metformin and sulfonylureas, which may reflect the financial burden of novel antihyperglycemic medications or the familiarity of prescribers and patients with well-established therapies as potential barriers to the consistent use of GLP-1 RAs and SGLT2i. The use of these drug classes in individuals with type 2 diabetes and established ASCVD has been suggested as a quality measure by Pharmacy Quality Alliance.^19^ The large attrition and inconsistent use reported in the present study has major implications on the practical definition of such quality measures, as initial prescription of these medications may not necessarily translate into their actual use by patients in the long term. As these medications become available for broader indications, such as obesity pharmacotherapy for GLP-1RAs^20,21^ and heart failure with preserved ejection fraction and CKD for SGLT-2i,^22-25^ addressing the utilization and persistence obstacles for their established indications grows increasingly important.

There are certain additional insights from these results that merit discussion. On average, higher income was associated with higher prescription fills of both medications among new initiators. Moreover, patients with commercial health insurance plans were more likely than Medicare Advantage beneficiaries to fill prescriptions after starting treatment. Notably, the difference between commercially insured and Medicare Advantage beneficiaries remained significant after adjusting for income, suggesting challenges with the Medicare Advantage plan design to facilitate affordable drug coverage. In 2019, median out of pocket costs for SGLT-2i and GLP-1 RAs in individuals with Medicare Advantage and Part D were estimated to range from $1,000 to $2,000 per year, depending on the prescribed agent and plan details.^26^ Our findings may reflect challenges with affordability with these agents, which continues to be a major obstacle to expand their use even among insured individuals.

Our study has limitations that merit consideration. First, our data does not represent all payers, limiting the generalizability of the present observations to the general population. Nevertheless, Optum Labs’ de-identified administrative claims data represents a diverse mixture of ages, races, and geographic regions across the US. Second, we were unable to account for insurance coverage differences for individual patients, manifested in plan-specific deductible thresholds, copay tiers, and cost sharing expectations, which may have implications for medication prescription and long-term adherence. Third, our dataset did not include uninsured individuals who are less likely to have access to these agents. Therefore, both the initial prescription and longitudinal fill rates for these agents in the general population are likely to be overestimated in our study. Fourth, we did not have access to the measures of glycemic control and drug side effects, which can potentially affect continuation of therapy among initiators of antihyperglycemic medications. However, we attempted to account for the severity of diabetes by including DCSI and CCI scores in our models. Fifth, all comorbidities were determined using claim-based indicators during the baseline period, which may underrepresent the true prevalence of comorbidities (e.g., diabetic nephropathy) in comparison with lab-based diagnostic methods; our assessment of ASCVD was limited to established disease, and not high ASCVD risk, which depending on the specific guideline or society recommendation, may be considered another indication for GLP-1 RAs and SGLT-2i. Finally, our proxies of income were derived at the zip-code level, not the individual level, and we were unable to account for other key social determinants of health such as occupation or education level.

## CONCLUSION

In this large nationwide US study on the real-world use of GLP-1 RAs and SGLT-2i, these medications were used in only 1 in 8 individuals with type 2 diabetes and indications in 2018-2020, with 50% drop off in prescription fills one year after initiation of therapy. The low frequency and inconsistent use of these evidence-based medications represents a challenge to realizing their longitudinal health outcomes benefits in a period of expanding indications of their use.

## Data Availability

The data are proprietary and are not available for open sharing due to restrictions on our data use agreement

## FUNDING

The study was funded by Research & Development at UnitedHealth Group, and the authors Callahan Clark, Abraham Reddy, and Samuel Amodeo are full-time employees of the UnitedHealth Group. These authors played an active role in all aspects of the development of the study, including design and conduct of the study; collection, management, analysis, and interpretation of the data; preparation, review, or approval of the manuscript; and decision to submit the manuscript for publication. The study was supported by the National Heart, Lung, and Blood Institute of the National Institutes of Health under award, 1K23HL153775 to Dr. Khera. The funder had no role in the design and conduct of the study; collection, management, analysis, and interpretation of the data; preparation, review, or approval of the manuscript; and decision to submit the manuscript for publication.

## DISCLOSURES

Callahan Clark, Abraham Reddy, and Samuel Amodeo are full time employees of UnitedHealth Group and own stock in the company. Marc Suchard reports grants from the National Science Foundation and the National Institutes of Health during the conduct of the study and grants from Janssen Research and Development outside the submitted work. Darren McGuire reports personal fees from Boehringer Ingelheim, Sanofi US, Merck & Co., Merck Sharp and Dohme Corp., Eli Lilly and Company, NovoNordisk, AstraZeneca, Lexicon Pharmaceuticals, Eisai, Pfizer, Metavant, Applied Therapeutics, Afimmune, Bayer, CSL Behring and Esperion. Zhenqiu Lin works under contract with the Centers for Medicare & Medicaid Services to develop quality measures. Silvio Inzucchi has received honoraria or consultancy fees from AstraZeneca, Boehringer Ingelheim, Novo Nordisk, Lexicon, Merck, vTv Therapeutics, Esperion, and Abbott. Evangelos Oikonomou and Rohan Khera are coinventors of US Provisional Patent Application No. 63/177,117, Methods for neighborhood phenomapping for clinical trials and are co-founders of Evidence2Health, a precision health and digital health analytics platform. The remaining authors report no potential conflicts of interest.

## Notes

### Author Declarations

The UnitedHealth Group Office of Human Research and the Yale Institutional Review Board exempted this study from review and waived informed consent as the study was limited to retrospective analyses of de-identified data and compliant with the Health Insurance Portability and Accountability Act (HIPAA) of 1996.

