## Supplemental tables and figures for "Patterns of Medication Use and Prescription Fills for Cardioprotective Anti-Hyperglycemic Agents in the United States"

### eTable 1. Inclusion criteria to select study population

| |
|---|
| **Inclusion Criteria:** |
| Individuals 18 years of age and older, AND |
| 36 months of continuous enrollment between January 2018 to December 2020, AND |
| Two or more claims with primary or secondary diagnosis of type 2 diabetes > 90 days apart |

**eTable 2. Medication list with individual agents in each drug class.** Abbreviations: GLP-1; glucagon-like peptide-1; SGLT-2i, sodium glucose cotransporter-2 inhibitor; DPP-4, dipeptidyl peptidase-4.

| **Drug Class** | **Agents** |
|---|---|
| **GLP-1 receptor agonists** | Albiglutide, Dulaglutide, Exenatide, Insulin Degludec-Liraglutide, Insulin Glargine-Lixisenatide, Liraglutide, Lixisenatide, Semaglutide |
| **SGLT-2i** | Canagliflozin, Canagliflozin-Metformin, Dapagliflozin, Dapagliflozin-Metformin, Dapagliflozin-Saxagliptin, Empagliflozin, Empagliflozin-Linagliptin, Empagliflozin-Metformin, Ertugliflozin, rtugliflozin-Metformin, Ertugliflozin-Sitagliptin |
| **Biguanides** | Alogliptin-Metformin, Canagliflozin-Metformin, Dapagliflozin-Metformin, Empagliflozin-Metformin, Ertugliflozin-Metformin, Glipizide-Metformin, Glyburide-Metformin, Linagliptin-Metformin, Metformin, Pioglitazone-Metformin, Repaglinide-Metformin, Rosiglitazone-Metformin, Saxagliptin-Metformin, Sitagliptin-Metformin |
| **Sulfonylureas** | Acetohexamide, Chlorpropamide, Glimepiride, Glipizide, Glipizide-Metformin, Glyburide, Glyburide, Glyburide-Metformin, Pioglitazone-Glimepiride, Rosiglitazone-Glimepiride, Tolazamide, Tolbutamide, Tolbutamide |
| **Thiazolidinediones** | Alogliptin-Pioglitazone, Pioglitazone, Pioglitazone-Glimepiride, Pioglitazone-Metformin, Rosiglitazone, Rosiglitazone-Glimepiride, Rosiglitazone-Metformin, Troglitazone |
| **DPP-4 inhibitors** | Alogliptin, Alogliptin-Metformin, Alogliptin-Pioglitazone, Dapagliflozin-Saxagliptin, Empagliflozin-Linagliptin, Ertugliflozin-Sitagliptin, Linagliptin, Linagliptin-Metformin, Saxagliptin, Saxagliptin-Metformin, Sitagliptin, Sitagliptin-Metformin, Sitagliptin-Simvastatin |
| **Meglitinides** | Repaglinide, Nateglinide |
| **Long- and Intermediate-Acting Insulins** | Insulin Degludec, Insulin Degludec-Liraglutide, Insulin Detemir, Insulin Glargine, Insulin Glargine-Lixisenatide, Insulin Isophane, Insulin Isophane (Pork), Insulin NPH (Human), Insulin Zinc, Insulin Zinc (Human), Insulin Zinc (Pork), Insulin Zinc Extended (Human) |
| **Short and Rapid-Acting Insulins** | Insulin Aspart, Insulin Aspart (with Niacinamide), Insulin Aspart Protamine & Aspart (Human), Insulin Glulisine, Insulin Lispro, Insulin Lispro Protamine & Lispro, Insulin Lispro-AABC, Insulin NPH Isophane-Regular (Human), Insulin Regular, Insulin Regular (Human), Insulin Regular (Human) Buffered, Insulin Regular (Human) in sodium chloride, Insulin Regular (Pork) |
| **Oral anticoagulants** | Apixaban, Rivaroxaban, Betrixaban, Edoxaban, Dabigatran, Warfarin |
| **Statins** | Atorvastatin, Amlodipine-Atorvastatin, Simvastatin, Niacin-Simvastatin, Pitavastatin, Lovastatin, Niacin-Lovastatin, Fluvastatin, Pravastatin, Rosuvastatin |
| **Angiotensin-Converting Enzyme Inhibitors** | Benazepril, Benazepril-Hydrochlorothiazide, Captopril, Captopril-Hydrochlorothiazide, Enalapril, Enalapril-Hydrochlorothiazide, Fosinopril, Fosinopril-Hydrochlorothiazide, Lisinopril, Lisinopril-Hydrochlorothiazide, Moexipril, Moexipril-Hydrochlorothiazide, Perindopril, |

| | |
|---|---|
| | Perindopril-Amlodipine, Quinapril, Quinapril-Hydrochlorothiazide, Ramipril, Trandolapril, Trandolapril-Verapamil |
| **Angiotensin II Receptor Antagonists** | Azilsartan, Azilsartan-Chlorthalidone, Candesartan, Candesartan-Hydrochlorothiazide, Eprosartan, Irbesartan, Irbesartan-Hydrochlorothiazide, Losartan, Losartan-Hydrochlorothiazide, Olmesartan, Olmesartan-Amlodipine-Hydrochlorothiazide, Olmesartan-Hydrochlorothiazide, Medoxomil, Sacubitril-Valsartan, Telmisartan, Telmisartan-Amlodipine, Telmisartan-Hydrochlorothiazide, Valsartan, Valsartan-Hydrochlorothiazide |
| **Beta-adrenergic Blocking Agents** | Acebutolol, Atenolol, Atenolol-Chlorthalidone, Betaxolol, Bisoprolol, Bisoprolol-Hydrochlorothiazide, Carvedilol, Labetalol, Metoprolol Succinate, Metoprolol Tartrate, Metoprolol-Hydrochlorothiazide, Nadolol, Nadolol-Bendroflumethiazide, Nebivolol, Nebivolol-Valsartan, Pindolol, Propranolol, Propranolol-Hydrochlorothiazide, |

**eTable 3. Eligibility and exclusion criteria for GLP-1RAs and SGLT-2i.** Clinical classifications software refined (CSSR) and ICD-10 codes were used to determine eligibility and exclusion criteria. Abbreviations: GLP-1RA, glucagon-like peptide-1 receptor agonist; SGLT-2i, sodium glucose cotransporter-2 inhibitor.

| |
|---|
| **Eligibility Criteria for GLP-1RAs:**<br><br>Atherosclerotic Cardiovascular Disease: Defined as<br><br>Acute myocardial infarction (CIR009), or<br><br>Complications of acute myocardial infarctions (CIR010), or<br><br>Cerebral infarction (CIR020), or<br><br>Occlusion or stenosis of precerebral or cerebral arteries without infarction (CIR023), or<br><br>Peripheral and visceral vascular disease (CIR026) |
| **Exclusion Criteria for GLP-1RAs:**<br><br>Medullary Thyroid Carcinoma: Proxied by any primary diagnosis for thyroid cancer or any claim related to personal history of malignant neoplasm of thyroid (C73%, Z85.850)<br><br>Multiple Endocrine Neoplasia Syndrome Type 2: Proxied by any primary diagnosis for multiple endorcine neoplasia syndrome type 2-A, 2-B, or unspecified (E31.20, E31.22, E31.23) |
| **Eligibility Criteria for SGLT-2i:**<br><br>Atherosclerotic Cardiovascular Disease: As defined above<br><br>Heart Failure (CIR019)<br><br>Diabetic Nephropathy (GEN003) |
| **Exclusion Criteria for SGLT-2i:**<br><br>Severe Renal Impairment: Proxied by any primary diagnosis of chronic kidney disease stage IV-V, end stage kidney disease, or dialysis-dependence (N18.4, N18.5, N18.6, Z99.2) |

**eTable 4. Demographic and clinical characteristics of individuals receiving GLP-1RAs and SGLT-2i.** Data represent mean (SD) for age, median (IQR) for neighborhood income, and number (percent) for other variables. Abbreviations: GLP-1RA; glucagon-like peptide-1 receptor agonist; SGLT-2i, sodium glucose cotransporter-2 inhibitor; DPP-4, dipeptidyl peptidase-4; ACEI, angiotensin converting enzyme inhibitor; ARB, angiotensin receptor II blocker; DCSI, diabetes complications and severity index; ASCVD, atherosclerotic cardiovascular disease; CKD, chronic kidney disease; DKD, diabetic kidney disease.

| | GLP-1RA | | SGLT-2i | |
|---|---|---|---|---|
| | **User** | **Non-user** | **User** | **Non-user** |
| **Overall** | 46,697 | 540,960 | 37,311 | 550,346 |
| **Age (year)** | 68.38 (9.93) | 73.30 (8.98) | 68.5 (9.95) | 73.24 (9.01) |
| **Female Sex** | 25,811 (55.3) | 285,031 (52.7) | 17,116 (45.9) | 293,726 (53.4) |
| **Health Insurance** | | | | |
| **Commercial** | 9,285 (19.8) | 55,455 (10.2) | 9,289 (24.9) | 55,449 (10.1) |
| **Medicare Advantage** | 37,412(80.2) | 485,505(89.8) | 28,020(75.1) | 494,897(89.9) |
| **Neighborhood Income ($)** | 53,317 [45,988-62,535] | 53,317 [46,338-62,553] | 53,742 [46,338-62,931] | 53,317 [46,338-62,553] |
| **Medications** | | | | |
| **Biguanides** | 30,660 (65.7) | 325,555 (60.2) | 26,452 (70.9) | 329,763 (59.9) |
| **Sulfonylureas** | 16,017 (34.3) | 151,652 (20.0) | 14,570 (39.1) | 153,099 (27.8) |
| **Thiazolidinediones** | 4,560 (9.8) | 31,509 (5.8) | 4,150 (11.1) | 31,919 (5.8) |
| **DPP-4 Inhibitors** | 7,448 (15.9) | 69,475 (12.8) | 10,902 (29.2) | 66,021 (12.0) |
| **Meglitinides** | 705 (1.5) | 4,736 (0.9) | 565 (1.5) | 4,876 (0.9) |
| **Insulin Long-acting** | 21,235 (45.5) | 94,098 (17.4) | 11,386 (30.5) | 103,947 (18.9) |
| **Insulin Short-acting** | 11,155 (23.9) | 59,716 (11.0) | 5,739 (15.4) | 65,132 (11.8) |
| **Statin** | 37,888 (81.1) | 397,230 (73.4) | 30,413 (81.5) | 404,705 (73.5) |
| **ACEI** | 20,959 (44.9) | 226,923 (41.9) | 17,083 (45.8) | 230,799 (41.9) |
| **ARB** | 17,623 (37.7) | 173,979 (32.2) | 13,252 (35.5) | 178,350 (32.4) |
| **Beta Blocker** | 21,010 (45.0) | 230,736 (42.7) | 15,141 (40.6) | 236,605 (43.0) |
| **Oral Anticoagulant** | 4,381 (9.4) | 54,321 (10.0) | 3,059 (8.2) | 55,643 (10.1) |
| **DCSI Score** | 2 (1.87) | 1.87 (1.83) | 1.65 (1.69) | 1.90 (1.84) |
| **Cardiovascular Disease** | 17,059 (36.5) | 212,457 (39.3) | 12,750 (34.2) | 216,766 (39.4) |
| **Cerebrovascular Disease** | 4,044 (8.7) | 54,200 (10.0) | 2,923 (7.8) | 55,321 (10.1) |
| **Metabolic Disease** | 1,949 (4.2) | 16,082 (3.0) | 1,283 (3.4) | 16,748 (3.0) |
| **Neuropathy** | 17,422 (37.3) | 152,133 (28.1) | 11,591 (31.1) | 157,964 (28.7) |
| **Peripheral Vascular Disease** | 8,650 (18.5) | 97,030 (17.9) | 6,080 (16.3) | 99,600 (18.1) |
| **Retinopathy** | 10,036 (21.5) | 89,978 (16.6) | 6,914 (18.5) | 93,100 (16.9) |
| **Nephropathy** | 13,946 (29.9) | 145,490 (26.9) | 7,308 (19.6) | 152,128 (27.6) |
| **ASCVD** | 15,757 (33.7) | 194,226 (35.9) | 12,018 (32.2) | 197,965 (36.0) |
| **Heart Failure** | 5,220 (11.2) | 63,286 (11.7) | 2,973 (8.0) | 65,533 (11.9) |
| **CKD** | 8,689 (18.6) | 91,026 (16.8) | 3,830 (10.3) | 95,885 (17.4) |
| **DKD** | 13,946 (29.9) | 145,490 (26.9) | 7,308 (19.6) | 152,128 (27.6) |

**eFigure 1. Trends of new prescriptions for GLP-1 RAs and SGLT-2i during 2019-2020.** Data represent number of new prescriptions for GLP-1 RAs and SGLT2i per 10k eligible individuals during the study period from 2019 through 2020. Abbreviations: GLP-1; glucagon-like peptide-1 receptor agonist; SGLT-2, sodium glucose cotransporter-2 inhibitor.

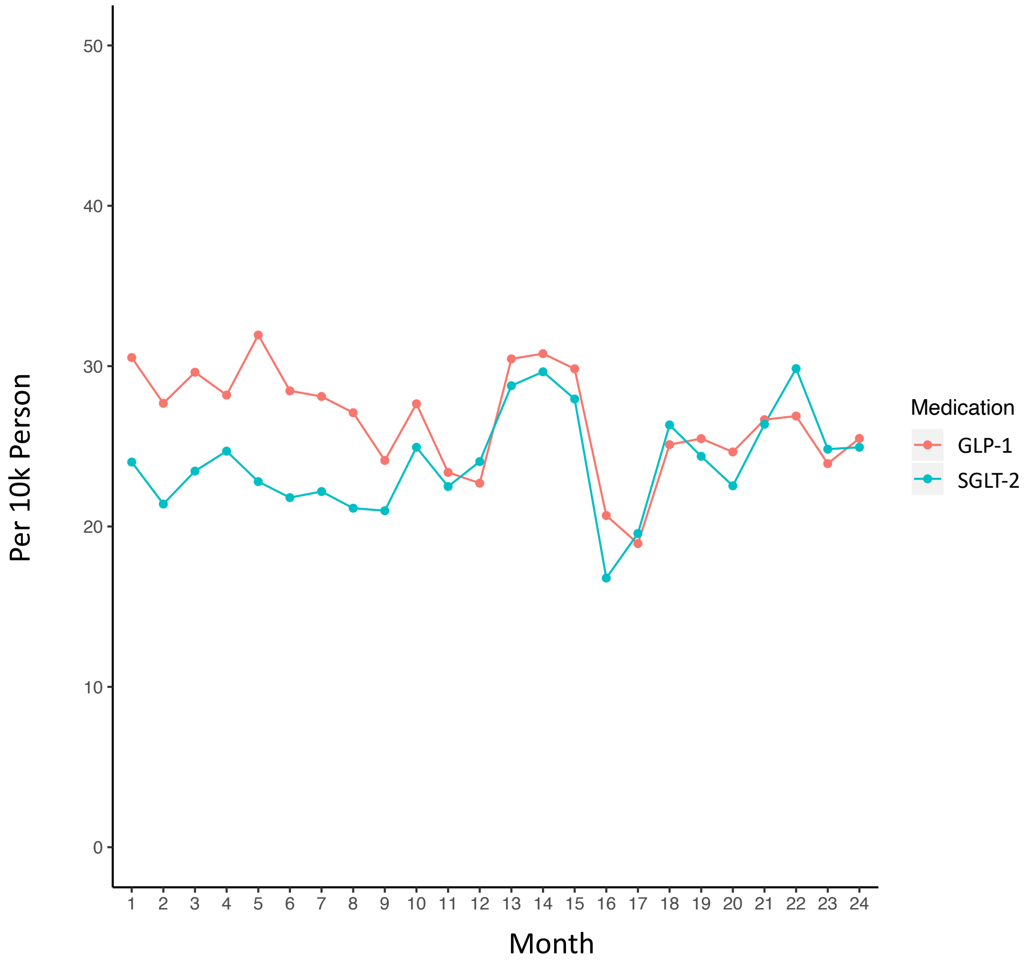

**eFigure 2. Monthly fill rates for GLP-1 RA and SGLT-2i prescriptions among new initiators across patient subgroups in 2019-2020.** Data represent monthly fill rates based on the proportion of days covered among new initiators of each drug class for 12 months after initiating treatment. Abbreviations: GLP-1; glucagon-like peptide-1 receptor agonist; SGLT-2, sodium glucose cotransporter-2 inhibitor; ASCVD, atherosclerotic cardiovascular disease.

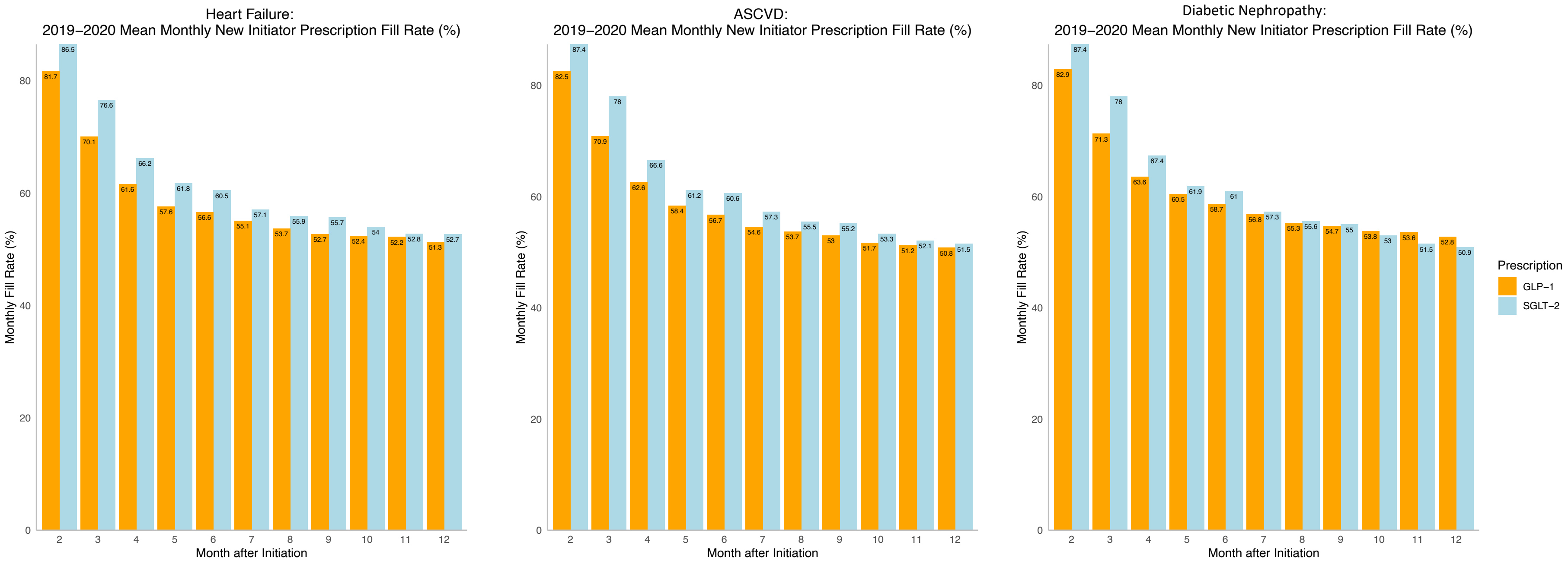

**eFigure 3. Monthly fill rates for GLP-1 RA and SGLT2i prescriptions among new initiators with Medicare Advantage and commercial insurance plans.** Data represent monthly fill rates based on the proportion of days covered among new initiators of each drug class for 12 months after initiating treatment. Abbreviations: GLP-1; glucagon-like peptide-1 receptor agonist; SGLT-2, sodium glucose cotransporter-2 inhibitor.

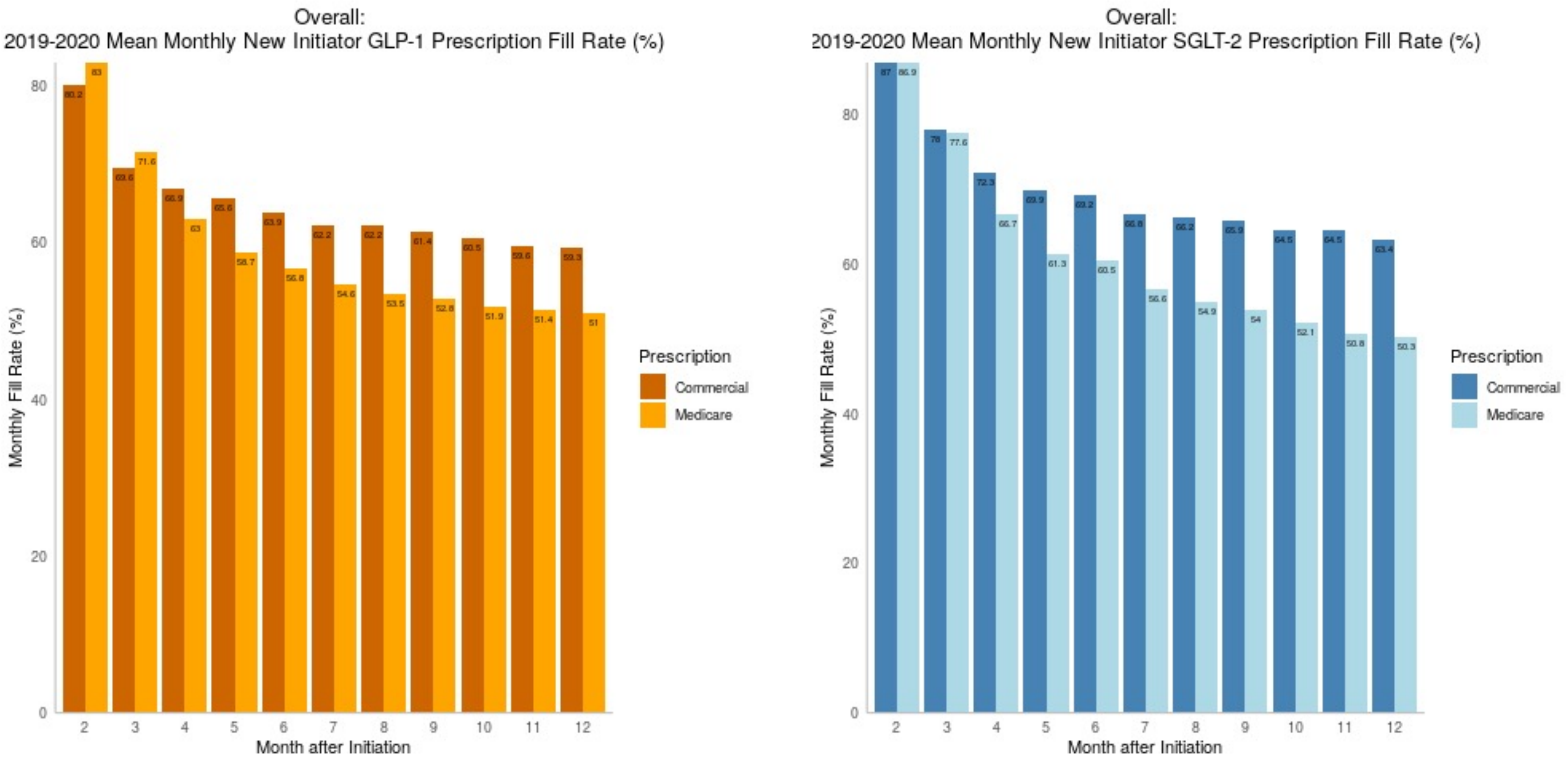

**eFigure 4. Monthly fill rates for metformin and sulfonylureas among new initiators in 2019-2020.** Data represent monthly fill rates based on the proportion of days covered among new initiators of each drug class for 12 months after initiating treatment.

(A) Monthly Fill Rates for Metformin

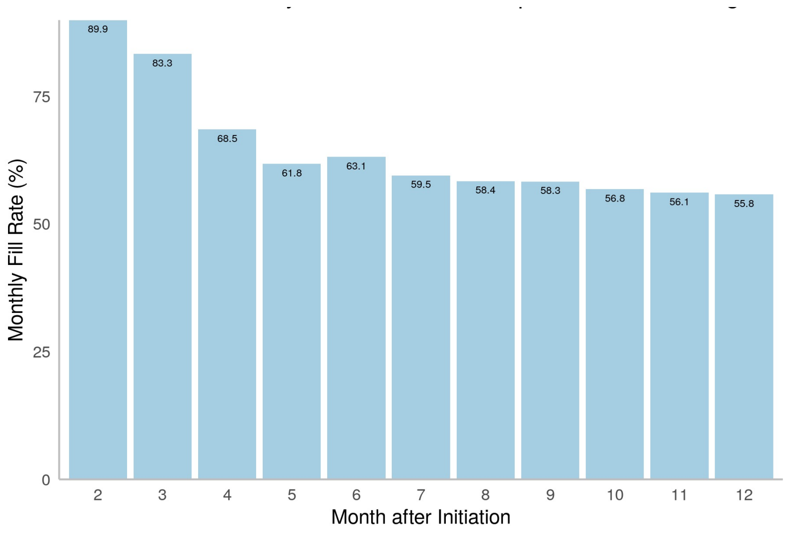


(B) Monthly Fill Rates for Sulfonylureas

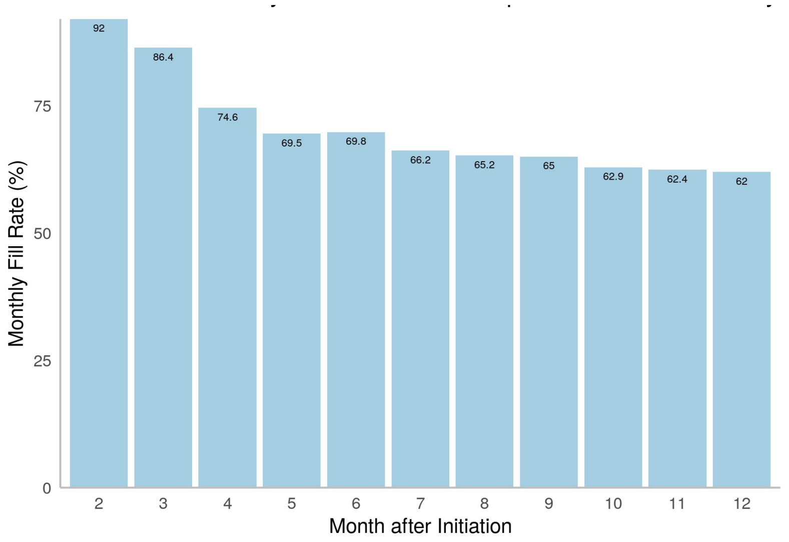
